# “They don’t know how to live with a child with these conditions, they can’t understand…”: The lived experiences of parenting a child with a genetic neurodevelopmental disorder

**DOI:** 10.1101/2024.07.24.24310802

**Authors:** Karen J. Low, Georgia Treneman-Evans, Sarah L Wynn, GenROC Study Consortium, Jenny Ingram

## Abstract

**Background:** A genetic neurodevelopmental diagnosis (GND) impacts all aspects of a child and family’s life. GNDs are rare; most have limited natural history data. We aimed to understand parents’ experiences around data acquisition about their child’s GNDs which can help inform clinical practice.

**Design and participants:** This analysis is part of the UK multicentre GenROC study. We conducted 17 semi-structured interviews with parents of children with GNDs (aged 0-15 years). Data were analysed following the principles of thematic analysis.

**Results:** Five main themes are reported:

*Impact on the family around a genetic diagnosis:* Distress results from diagnosis wait, the act of receiving it, associated irreversibility (loss of hope) and family/reproductive implications. GNDs and Uncertainty: Lack of data and rareness causes uncertainty for the future.

*Relationships with health professionals:* Positive where parents are empowered and feel part of the team; Negative –parents feel not heard/believed or lack of expertise/understanding.

*Parent mental health:* GNDs can be a significant burden to family life. Need for advocating for services is a negative impact. Isolation through rareness is a factor – this can be helped by support networks which mostly consist of gene specific Facebook groups.

*Development of positive parent identities:* including that of advocate, professional and educator.

**Conclusions:** GNDs represent a major challenge for families, clinicians and service providers. Distressed parents are struggling to cope with challenges and suffer poor mental health. Psychosocial support, better signposting, and health professional education may help.

**Patient contribution:** PPI group contributed to topic guide development and commented on findings.

## Background

More children are being diagnosed with rare genetic neurodevelopmental disorders (GNDs) than ever before in the UK, often within non-specialist settings, and sometimes with the diagnosis being received at a very early age. Falling cost of Whole Genome Sequencing(WGS) has allowed rapid implementation in England. A child is deemed eligible for testing based on nationally specified criteria.

Some studies (1–4) have assessed the impact of a genetic diagnosis on families. However, there are limited data documenting parents’ and carers’ (from here on collectively referred to as parents) views on their experience and levels of support as they then go on to navigate the landscape of having a child with a rare condition. Data that do exist are largely limited to populations where testing was purely undertaken by specialists. Insight into parent’s experiences of receiving genomic diagnosis alongside their lived experience is needed to inform policy and clinical practice as WGS continues to be implemented widely.

We aimed to understand the lived experience of parents of children with GNDs to identify the needs, obstacles and barriers for families. Our primary research question was to understand what information is currently available, what is missing for parents of GNDs and how are they adapting to address this information gap. These results may inform practice and underpin future recommendations for improving family support and clinical practice.

## Methods

### Study design

This qualitative study is part of the national multi-centre study “Improving the clinical care for Genetic Rare disease: Observational Cohort Study” (GenROC: REC22/EM/0274) (5). We aimed to identify the information gaps for parents and understand the lived experience of parenting a child with a GND.

To understand their experience, the first author undertook semi-structured interviews with parents of children with known GNDs aged 0-15 years. Interviews were based on a semi-structured topic guide that had undergone ethical approval as part of the wider approval process for the GenROC study (6). All interviews took place online with participants in their homes using the video conferencing platform, Zoom. One participant had their child present. The topic guide was informed by the relevant literature and developed with professionals and stakeholders in the area of rare disease as well as the study’s Patient Participant Involvement (PPI) group. The interviewer was a senior clinician who has had training in qualitative research methods and no prior contact with the participants.

The topic guide follows a pre-defined structure but allowed additional questions and emerging issues in the context of GNDs. This guide includes [1] an introduction of the study and interviewees, then focussed on [2] the families’ pathway leading up to and during the diagnosis, [3] daily living with a GND and its impact on navigating healthcare, [4] what information families have been missing and how they have looked for this information and accessed it including areas of support and [5] if and how they utilise digital health and social media in this respect and [6] a chance for parents to discuss anything else that they felt was important(see supplementary file 1).

### Participants and sampling

Inclusion and exclusion criteria for participation in the interview were as for enrolment in-to GenROC (6).

Parents completed consent to being participating in the GenROC study and they were given an option to consent to being contacted about an interview. 91% of parents agreed to this option. We purposively sampled to ensure maximum variation across the following categories 1) postcode Indices of Multiple Deprivation(IMD) centile; 2) type of genetic disorders (figure 1) 3) UK geography; 4) ethnicity; 5) children of differing ages 6) fathers were actively recruited (mothers were also invited); 7) foster and adoptive parents; 8) parents advocates, leads of support groups or charities.

**Figure 1:**
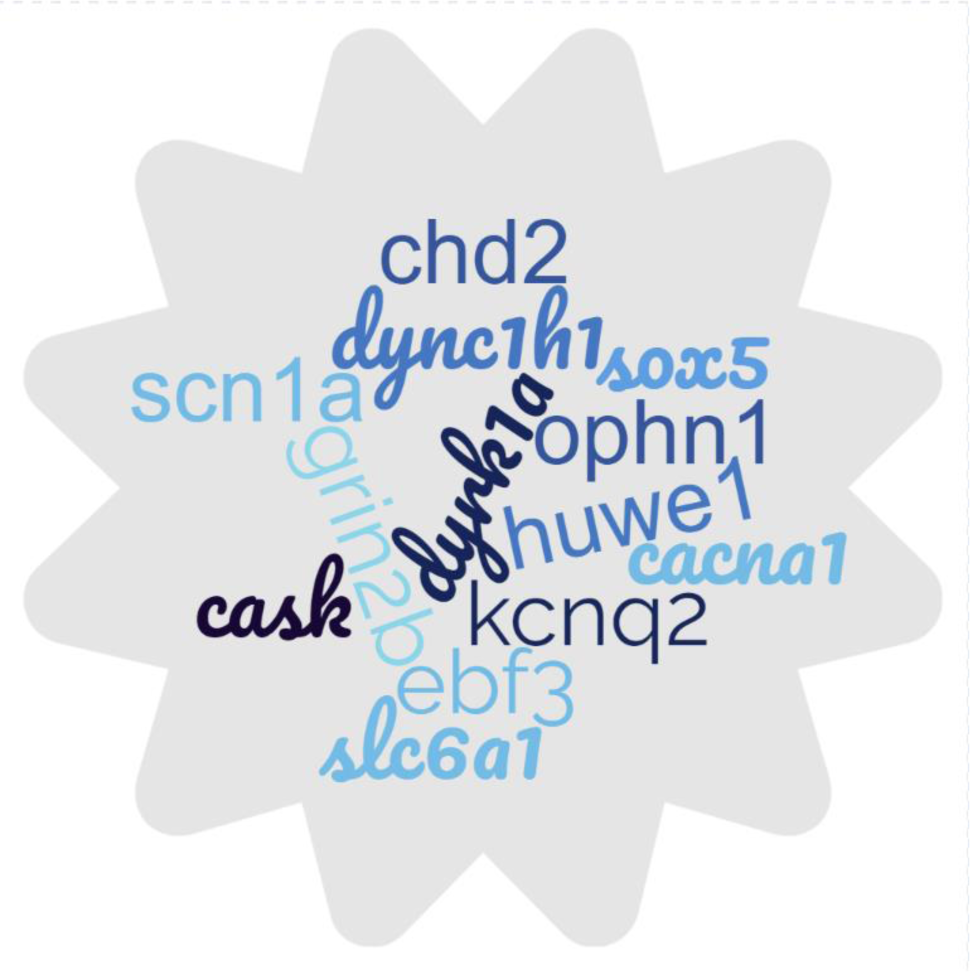
Word cloud of genetic conditions represented in participant sample.

Parents were invited to take part by email. They were informed about the study using a Parent Information Leaflet which had undergone ethical approval as part of the wider GenROC study (6). Parents were required to complete an interview specific consent form prior to interview. Appointments dates and times for the interview were agreed via email. No participants refused to participate nor did any drop out.

### Data collection

The interviews were conducted between November 2023 and March 2024. Interviews were conducted by the lead author who is chief investigator of the GenROC study. All interviews were recorded, transcribed verbatim and anonymised and combined with interviewer’s field notes for analysis. Interviews continued until no new themes were being generated. Transcriptions were imported into NVIVO qualitative data management software (Release 1.71) for coding and data management. No repeat interviews were undertaken.

### Data analysis

Data were analysed following the principles of thematic analysis (7). An iterative approach was used to developed an initial codebook informed by the aims of the study (deductive component). The first 5 transcripts were coded by the first author and additional codes that were not covered by the initial codebook were then added (inductive component). 20% of the transcripts were double coded by the second author, an experienced qualitative researcher. All discrepancies were reconciled through consensus. NVIVO software was used to group codes and these were refined following discussions between the authors. Methodological oversight was provided by the final author. Reporting of data has been performed according to COREQ guidelines (8).

## Results

All sampled participants accepted the invitation. 13 interviews were conducted in total with a total of 17 parents whose characteristics are summarised in Table 1. Interviews lasted an average of 46 minutes with the shortest being 27 minutes (single parent interview) and the longest being 58 minutes (2 parent interview). Five themes were identified: *Impact on the family around a genetic diagnosis; GNDs and Uncertainty; Relationships with health professionals (negative and positive); Parent mental health*; *Development of positive parent identities* as shown in Table 2. Themes are presented with illustrative quotes in tables 3-8 and identified by an anonymised number and indicating whether ‘mother or father’.

### Theme 1: “Please just tell me what’s wrong with my baby?” And they were like, “We don’t know. We need these results back.” : Impact on the family around receiving a genetic diagnosis (Table 3)

#### Distress leading up to a diagnosis

Parents often describe having a gut feeling that something was wrong from an early stage. For some, this may have been in association with medical concerns such as scan abnormalities or reduced movements in pregnancy. For others this was once their baby was born when they failed to meet expected milestones or where they were displaying unusual symptoms or behaviours.

It often takes multiple visits and involvement of many healthcare professionals before a child was recognised as possibly having a genetic diagnosis. This can be very distressing to parents, leave them feeling unheard, anxious and alone in their concerns. (Q1). Parents gut feeling that something was wrong leads them to persist to press for answers despite being told their child is healthy, often by more than one professional.

Some parents start researching possible genetic conditions that may fit with their child’s physical features and medical conditions prior to having genetic testing. They also describe how their education and work background sometimes impacts positively or negatively on health professional attitudes with examples being a biology teacher and a shop worker respectively. (Q2)

In some cases parents reported that they felt that a genetic test was only agreed to a consequence of the amount of time the parent had spent complaining and raising concerns about their child.

Parents universally describe a difficult period of waiting for the results. Additionally, parents are often informed that a result is available but they have to wait for an appointment to discuss this result which exacerbates their distress and anxiety. One parent was told a result was available but the earliest appointment was in three weeks’ time. She was so desperate for the result that on the day of the appointment she went and sat in the waiting room 4 hours early in the hope she might be seen sooner. (Q3)

#### Receiving a genetic diagnosis

Parents describe receiving devastating information which was sometimes delivered in a very casual or informal manner. (Q4) Professionals deliver diagnoses in differing settings and formats. Sometimes this was only over the phone or video call (noting that some of those interviewed received diagnoses during COVID). Telephone conversations do not always allow parents sufficient time to digest the information. Complicated concepts such as “misfiring potassium channels” are not always understood over the phone which can impact parents own research into the condition later on.

Some parents were given the diagnosis initially by a paediatrician but then told they needed to be referred to a geneticist for further discussion. This additional wait heightens distress to parents who reported that they would have found it easier without a time lapse.

Parents also describe the difficulty in receiving a diagnosis and complex information during an appointment in which they are also expected to manage their child’s needs; one parent described the appointment as a marathon (Q5)

Parents all report that the genetic diagnosis had been helpful to them and their child. The reasons for this include removing guilt around something they may have done to cause their child’s challenges. It also enables them to obtain support for their child including benefits such as Disability Living Allowance or agreement for additional educational support in the form of an Educational Health Care Plan (EHCP). It can inform the clinicians regarding tailored treatment and monitoring plans for their child. Families can access condition specific support groups. It also can enable some idea of what to expect for their child in the future.

Parents report that prior to having a confirmed diagnosis their child’s complexity can be dismissed as parental anxiety. In contrast the diagnosis reduces the burden to some degree on parents to explain the extent of a child’s problems to professionals as the genetic diagnosis is “all-encompassing.” The diagnostic conversation is commonly followed up with a summary letter. Some parents reported frustration around the lack of detail provided to them. This was most specifically related to not being provided with their child’s specific genetic variant (Q6).

#### Implications for the wider family

A GND could have implications for reproductive decision making, but parents reported difficulty making decisions because the recurrence risks quoted to them were unclear or differed between health professionals and the support groups/research. Following parental testing one family had been counselled that they had a low chance of having another affected child. They described the distress in the slow dawning realisation that their second child was looking increasingly likely to have the same GND. (Q7) Parents were grateful that if their child goes on to have children of their own they will be better prepared for that child having potential issues and this could reduce the distress that they had experienced.

### Theme 2: “Sometimes I sit and think maybe I wish he did just have Down’s syndrome, and then everyone knows it [rather] than having X [and] no one knowing it” : The impact of uncertainty, lack of data and “rareness”(Table4)

Some parents are anxious that having a genetic diagnosis can result in a child’s challenges being incorrectly attributed to that condition and not being correctly investigated. (Q8)

Detailed natural history data for GNDs are often limited which can lead to parental frustration and worry when their child develops a new problem. (Q9)

Parents noted that the pace of knowledge availability and development is very slow. The lack of data also means that some parents struggle to plan and manage expectations for their family and the future (Q10)

Rareness can be distressing and can feel overwhelming, especially when the numbers of children affected worldwide are very small. Some parents reported that in some ways they would prefer to have a child with a better-known genetic condition because then the burden of explanation wouldn’t be so high.

Parents realise quickly that the professionals who are giving them the diagnosis often know very little about the condition and simply direct them to Facebook support groups and patient information leaflets (if they are available). Professionals tell parents that there are too many rare conditions which means it is not possible for professionals to know very much about them in detail. As a result, they often feel they need to seek information out themselves which adds to their mental load. It can lead to parents perceiving a feeling of disinterest or disengagement from the professional. Due to the lack of expertise parents would like to have a named clinical expert for their child’s genetic condition, however this is very variable as some conditions do not have one and for many the “expert” is in another country. Parents clearly stated that they wanted a UK based named expert for their child’s condition who could coordinate care nationally.

Rareness means that everyday interactions with other people are harder due to general lack of awareness and understanding. This places an additional burden of explanation on parents to extended family, friends and wider community. Some describe the frustration of people trying to suggest they have lived experience when they do not. (Q11)

Parents find the genetic diagnosis helpful, but it can also have a negative impact on them emotionally. Some report the unfairness or “bad luck” of having a genetic condition. (Q12) Many parents spoke about the finality of a genetic diagnosis and how “it is for life” and can remove hope of a child getting better or being “normal”. (Q13)

### Theme 3: “You are made to feel that you are going mad” :Relationship with health professionals (Table 5)

**Table 5:** Relationship with health professionals.

| Quote number | Quote |
| --- | --- |
| 14 | Anyway he did phone, and I was all by myself, and he was very jubilant in that he'd been right. So it was a, "Yes I was right, I'm amazing, she has X," which obviously if you Google X only has a very short life expectancy." (R13F) |
| 15 | "..you'll go in, and instead of being supportive to you, they argue against you, and fight against you, and I'm just like just listen to me, I know my son, I've been with him two years now. I always end up being right don't I in the end?" (R8F) |
| 16 | "Yeah, and then they look at you and then sometimes shut you down and tell you, you don't know what you're talking about. That gene what you're talking about doesn't do that kind of thing, and you're having to fight, ..." (R2F) |
| 17 | "But I have been really, really disappointed in the doctor care that we have received, and it has been consistent gaslighting and diversion and refusal to provide service, consistent for three years." (R7F) |
| 18 | "They don't know how to live with a child with these conditions, and they don't know, they can't understand ... but you and your wife haven't slept for two weeks, and you've got some other issue, and you've got something else, and you've got a busy day at work, or something like that, and it's putting it all together in your life, but for them they'll just skip over a fact because it's they want to move onto the next thing." (R5M). |
| 19 | "we saw a better paediatrician luckily, who did spend an hour and a half with us the first appointment which now that go and see him fairly regularly I know that was a big chunk of his morning clinic that day, and he literally just let me talk, and he said, "Is there anything else? And is there anything else you're worried about?" (R10F) |
| 20 | Because when I spoke to [doctor] she's like, "Oh yes the [specific mutation] kiddos," and she's talking about it like everybody knows that's a hotspot, and no one had ever said that to me before, and I wasn't aware that actually although he had this super rare thing he's in a quite common part of the super rare thing, which is good and bad to hear I guess." (R10F) |
| 21 | "So it's really hard, but unfortunately that's the only place they do it, and unfortunately the only time they do the conference is July, so it's 40 degrees, super humid, not a great time to take a family on holidays to X. I looked at the flights the other day for three of us and it was £4,000 already to go next summer. So yeah we'll have to see. But it was really valuable." (R10F) |

Parents universally report distress regarding professional’s attitudes and behaviours. One behaviour included a professional demonstrating happiness over being correct with their diagnostic prowess despite the associated negative outcome of said condition. (Q14)

Parents describe that they are required to advocate for their child endlessly which can be extremely hard, tiring and upsetting. (Q15) A foster carer described that the fight was even harder for her because professionals questioned her concerns and implied that she didn’t know her child well enough yet.

Parents feel continuously doubted by professionals. There is an implied hierarchy with the professional automatically believing they should be *de facto* more expert than the parent resulting in difficult interactions when parents present new data or question the clinician’s opinion. (Q16) Professional egos are sometimes reported as another factor with parents perceiving that some professionals (particularly hospital consultants) don’t like to be told what to do by a parent.

Parents can also feel frustrated when they read that their child should be receiving a treatment or therapy but are not given this for their child. (Q17) Service pressures result in parents having to be more demanding than they would wish to be which is exhausting and busy clinical environments can lead to lack of empathy. (Q18)

Many parents did identify some health professionals as being an important source of support. Being listened to without judgement and being given time and space to talk was highly valued. (Q19) Having a good relationship with the child’s key paediatrician was reported as being an important support. Parents valued the opportunity to have an egalitarian dialogue with this professional and that information that they brought was welcomed.

Parents value an expert professional in their child’s genetic condition but for most this expert is based in another country. Being able to speak to that expert is extremely desirable and reduces anxiety. (Q20) This wish for access to the expertise means that parents may travel to attend the international family conference at considerable personal expense which would not be possible for many families and can be impossible with complex childcare needs and lack of a wider family network to help. (Q21)

### Theme 4: “Then like a period of mourning really. You have to let go of the future you thought was going to happen, and you adapt…you’re a parent to a severely disabled child, and it touches every area of everything you do” : Parent mental health (Table 6)

Many parents described that having a child with GND along with lack of support has resulted in a significant deterioration in their mental health. (Q19) Parents report significant distress due to the constant need to fight with professionals to access care and support for their child; in one case this battle led to a social services referral.

Many parents are assumed to be overanxious by professionals which can lead to them fighting harder for their child and negatively impacting their mental health. (Q20) Parents describe the burden of living with GND as all encompassing, and they experience a period of grief around the diagnosis. (Q21)

Managing children’s challenging or unsafe behaviour is particularly difficult for families and results in them being hampered in what activities they can achieve as a family unit. Parents need to be constantly vigilant and cannot get respite placing a huge demand on them daily. Parents can find the comparison with “normal” families very upsetting. (Q22)

### Theme 5: “Social media (Facebook) has been instrumental and key to feeling supported and making connections, and having people who very specifically understand your experience”: Coping strategies and factors that help parents (Table 7)

#### Shared lived experience and support groups

Shared lived experience with other families is the most highly reported coping strategy. Some parents describe the friendships that they have made through support groups much of which takes place via social media (Q23). When a child develops a new problem parents will use the group as a sounding board about the new concern and will use the responses to help them determine what course of action (if any) should be taken. (Q24)

Groups allow parents to assess for themselves where their child fits in the spectrum of the condition and what the future may hold in a way that isn’t possible using “dry” scientific literature. (Q25) Lack of natural history data and variability of phenotype expression leads parents to worry about what the future holds for their child. They look to support groups for examples of older individuals (Q26). Finding examples of individuals who have achieved certain milestones in education or independence provide parents with hope. (Q51)

Parents consult the groups to get practical information about toys, types of mobility support aids, and days out amongst others. Some parents in our study describe other parents in the group as “good friends” indicating the strength of bond that had developed and reported sending and receiving birthday cards and presents including when their child received gifts when admitted to hospital.

Whilst most of the groups interacted predominantly online, one UK group organised an annual family weekend at a holiday park, part funded by fundraising grants. A parent reported how beneficial these weekends were in terms of providing a support network including for other siblings. A big benefit for them was seeing other families who had gone on to make the choice to have another (unaffected) child which influenced their own family decisions. (Q27)

#### Parents as experts or professionals

Parents consistently described the desire and ultimately the need to become an expert in their child’s rare condition in order to navigate accessing appropriate health, education and support. Becoming an expert appears to have unexpected collateral benefits. Some parents, particularly those who have a scientific education or an interest in science, report an enjoyment associated with the genomic science of their child’s condition and with the process of researching it. For some parents, this also allows them to feel that they are actively doing something to help in the context of a situation in which they frequently feel helpless. Fathers were more frequently described in this way. (Q28)

A smaller number of parents evolved to become peer supporters and provide informal advice to less well-informed parents or parents of newly diagnosed children. This role as “advisor” provides purpose and self-esteem as well as the positive benefits of being able to help other families. (Q29) Some parents in our study have set up foundations and charities which opened up a whole new career for them. One of these mothers reported how important that was for her as she had had to give up her career to care for her child and how without this evolving role she would have felt that she had lost herself.

Foster parents are professionals who choose to look after children with challenging complex needs and describe themselves as “project managers.” They describe great fulfilment from their role both personally and professionally. Other parents describe an evolution from parent as expert to advocate and professional. Some have set up condition specific websites, foundations and charitable organisations dedicating considerable time and effort to this, often on a voluntary basis. They describe linking in with professional groups, attending conferences and making presentations in multiple settings, enabling them to continue to develop personally and professionally and giving them fulfilment in a role other than parent with considerable positive impact. (Q30)

#### Being aware of/ involved in research

Parents view the opportunity to take part in genetic research studies as a positive way to improve their chance of getting more data on their child’s condition. They also value the chance to help other families in the future and see this as a positive reason to engage with research. (Q31)

## Discussion

This study highlights the significant challenges faced by children with GNDs and their families. Parents experience considerable distress throughout all stages of their child’s care pathway, diagnostic journey, with accessing services and interacting with professionals. This distress can result in a deterioration in parents’ mental health. Optimising care to reduce distress factors could improve parents’ capacity to cope which could potentially have a positive impact on mental health.

The diagnostic wait is difficult for families (9). Access to genomic testing has improved in the last decade but many parents still describe a period of time in which they knew something was not right with their child but were dismissed by healthcare professionals or they could not access the appropriate service. Even in cases where a diagnosis was made at a young age, parents report the lag time leading up to that diagnosis as being distressing. Frustration and distress associated with the diagnostic odyssey has also been reported in parents interviewed as part of an undiagnosed diseases program (10). Nonetheless, there is clear evidence in the literature and from our findings that parents consider a diagnosis to be enormously beneficial as it gives a reason “why” and opens doors to services and support (1). Distress appears to be unavoidably related to the period of time where parents do not have an explanation or answers for their child’s symptoms. It may be that some of this distress is not avoidable as much of it relates to having a child who has problems, often unexpectedly. However, attempts to reduce waiting should be prioritised at all stages. This recommendation is exemplified by the findings of this study, where one parent sat and waited all day in an outpatient waiting room despite her appointment in the hope that they might see her sooner because she was there.

The lack of information in GNDs leads to distress and worry (3, 9) and this was apparent in the current findings. Most of the time it is not possible to accurately predict where their child would fall in the long-term compared to others raising anxiety and making it difficult to plan. Raising the profile of adults with these GNDs and allowing them to become mentors and advocates (as has been done for another rare disorder) (4) may be a mutually beneficial strategy going forward as more children who benefited from exome sequencing diagnoses grow up.

Parents may feel alone despite receiving a diagnosis (1). Shared lived experience and peer support is beneficial for many parents. This allows them to ask questions and find out information from other parents, including finding out about resources available for their children (11). This was echoed by some of the parents in our study who reported that peer support groups can give you useful insight into options they had been unaware of. Parents of GNDs can become socially isolated due to lack of understanding from society, complexity of their child’s needs (11, 12), due to worry and shame related to their child’s behaviour and people not being able to understand that their child had a complex underlying diagnosis. A virtual peer support group circumvents these barriers and reduces social isolation. The diagnosis discussion should include information about support groups and ideally signpost families to an appropriate group so that they are aware of these from an early stage.

A genetic diagnosis is not the same as any other paediatric diagnosis (13). A mixed methods study of the lived experience of parents of children with CDKL5 deficiency disorder cited grief, and an associated grieving journey, in the context of receiving the diagnosis as a nearly universal theme amongst parents (14). Our findings highlight that the “genetic-ness” of the diagnosis had very specific impacts on parents. For instance, parents felt that the genetic diagnosis means that the condition can’t be reversed and it’s not going to go away, and it removed hope of it getting better. These findings were echoed in Rare Minds 2024 survey (15). They found that 41% and 85% of respondents felt having or caring for someone with a rare condition had negatively impacted their family relationships and their mental health or emotional wellbeing respectively.

For some families and conditions, a genetic diagnosis removes hope. It is important that professionals recognise the way this differentiates this type of diagnosis from others. Psychotherapeutic counselling should be funded specifically for families with genetic disorders but currently is not available for most and very rarely offered (15). 96% of Rare Minds (15) respondents felt that it was important that mental health professionals had an understanding of rare conditions and how they can impact mental health. This agrees with our findings and we concur with the Rare Minds recommendation that anyone impacted by a rare condition should receive psychologically-informed ‘rare aware’ care(15).

A genetic diagnosis also has an impact for the wider family. The impacts for siblings and family life when having a child with any sort of complex medical condition or neuro-disability are well documented and include the stressors of managing emergencies, coordinating care, and advocating for your child and fighting the system (16). Lack of time, socio-legal difficulties and organisation problems are significant barriers (12). All parents in our study described some dissatisfaction (of varying proportions) with the healthcare system and professionals. They describe an endless battle to be believed and heard, to access the appropriate care, and to access specialist services. This impacts mental health and reduces their capacity to cope with additional challenges as well as negatively impacting patient-professional relationships. A centrally coordinated pathway for GNDs with centres of excellence may reduce this burden for families and ultimately improve clinical outcomes.

Parents develop considerable expertise in managing their child’s condition (17), and this can be both a burden and a source of self-fulfilment at the same time (11, 18). Our study shows that parents feel they become experts out of necessity due to the overwhelming need to know more, and to try and understand what the future holds. However, some parents take a more active role in becoming expert. For some fathers in our study, becoming an expert allowed them to feel that they were “doing something” active about their situation which allowed them to cope better with the challenges. Parents should also be signposted to further options for education explaining basic genetics to minimise their frustration in understanding the information.

It is important for professionals to provide parents with key information about the genetic diagnosis, including the variant details. Peer mentoring may provide a support mechanism in this regard particularly for parents who wish to become more active as a professional advocate (4). Parents desire a national named clinical expert for their child’s rare condition and consider this a priority. Implementation is challenging on a practical level given the number of different rare conditions, funding mechanisms and clinical set ups. This point should be considered as a medium-term strategic priority for rare disease.

### PPI reflections

Our PPI group reported our findings resonated very strongly with their lived experience (Q32,33,34). They recognised that professionals are doing their best in often difficult circumstances but hoped these findings would drive improved practice. One member reflected on the significant burden of being an expert in your child’s condition and the fight to be heard(Q35).

### Strengths and limitations

Our sample included both males and females. Some couples preferred to be interviewed together which may have resulted in one spouse having a more dominant voice. We interviewed couples and single parents as well as foster carers which provides unique insight. We purposively sampled to include a wide socioeconomic demographic and range of locations across the UK. We included a wide range of GNDs with some being more common and some less so(figure 1). We included parents who had become advocates and set up charities but also included parents who were not on Facebook at all and were not members of any groups. This spread has allowed for good sample diversity. Most, but not all, were white British so not all of our findings can necessarily be generalised to all groups. Our cohort only includes children with GNDs. This means that our findings may not be generalisable to all rare diseases. Due to the nature of the study some of our findings are likely to be relevant to wider patient groups such as those with chronic disease, other forms of disability and neurodiversity.

## Supporting information

Topic guide

## Conclusions and future directions

GNDs represent a major challenge for patients, parents, clinicians and service providers. Whilst genomic testing and consequently diagnoses are now more readily accessible and at a younger age, parents are universally distressed. Distressed parents are less able to cope with the challenges and suffer poor mental health. Based on our findings we have proposed practical recommendations for practice, outlined in figure 2.

**Figure 2:**
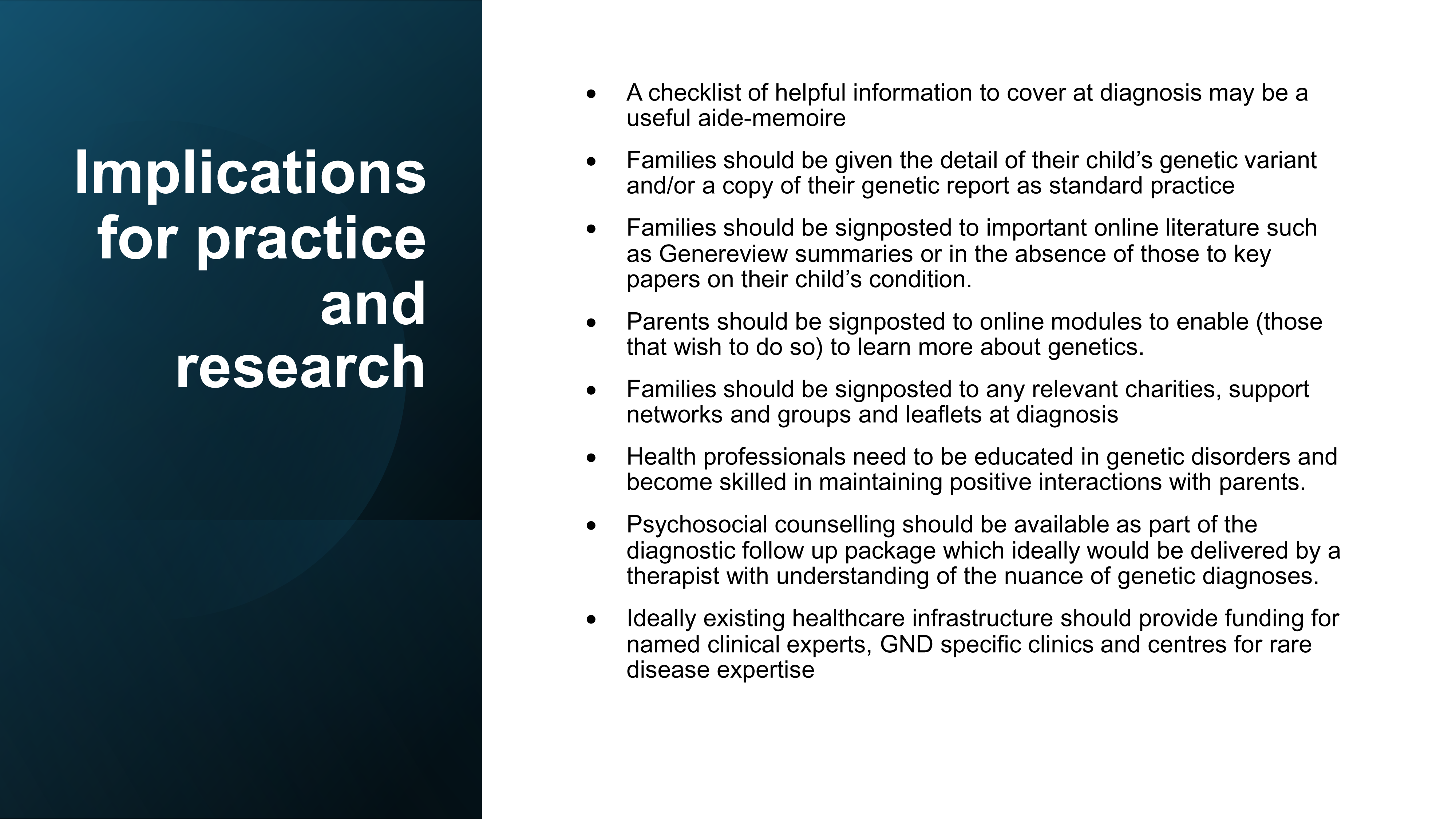
Recommendations for practice.

## Legends

Table 1: Table of participant sample character

Table 2: Summary of themes and subthemes

Table 3: Impact on the family of a genetic diagnosis

Table 4: Impact of uncertainty, lack of data and “rareness”

Table 6: Parent mental health

Table 7: Coping strategies and factors that help parents

Table 8: PPI group reflections

## Data availability

Clinically interpreted variants and associated phenotypes from the GenROC study are available through DECIPHER (https://www.deciphergenomics.org/). Transcripts from interviews are not available due to sensitivity of discussions and risk to vulnerable children.

## Acknowledgements

We thank all the parents in the GenROC study for contributing their views and the GenROC study PPI panel for their invaluable thoughts and guidance. The GenROC consortium is comprised multiple contributors from NHS sites across the UK. The consortium is comprised of individuals who have either acted as a local site PI or Associate PI or completed clinical proformas for GenROC. The list of individuals is as follows: Suzanne Alsters, Ruth Armstrong, Tazeen Ashraf, Queenstone Baker, Diana Baralle, Jonathen Berg, Marta Bertoli, Thomas Boddington, Moira Blyth, Catherine Breen, Helen Brittain, Lisa Bryson, Jenny Carmichael, Emma Clement, Tessa Coupar, Anna de Burca, Cristina Dias, Fleur Dijk, Abhijit Dixit, Alan Donaldson, Andrew Douglas, Jacqueline Eason, Fayadh Fauzi, Elaine Fletcher, Helen V. Firth, Nicola Foulds, Caroline Furnell, Andrew Fry, Laura Furness, Jennifer Gardner, Merrie Gowie, Rachel Harrison, Verity Hartill, Lizzie Harris, Eleanor Hay, Jenny Higgs, Simon Holden, Daniela Iancu, Rachel Irving, Vani Jain, Rosalyn Jewell, Gabriela Jones, Beckie Kaemba, Arveen Kamath, Ayse Nur Kavasoglu, Mira Kharbanda, Sophie King, Alison Kraus, Ajith Kumar, Katherine Lachlan, Neeta Lakhani, Wayne Lam, Anne Lampe, Abigail Lazenbury, Helen Leveret, Jessica Maiden, Alison Male, Alisdair McNeil. Ruth McGowan, Holly McHale, Catherine McWilliam, Jonathan Memish, Lara Menzies, Radwa Mohamed, Tara Montgomery, Oliver Murch, Michael Parker, Caroline Pottinger, Vijayalakshmi Ramakumaran, Ruth Richardson, Alison Ross, Claire Searle, Charles Shaw-Smith, Suresh Somarathi, Edward Steel, Helen Stewart, Kerra Templeton, Riya Tharakan, Madeline Tooley, Mohamed Wafik, Emma Wakeling, Elizabeth Wall, Amy Watford, Patricia Wells, Louise Wilson

## Funding statement

For the purpose of open access, the author has applied a CC-BY public copyright licence to any author accepted manuscript version arising from this submission. KL and GenROC is supported by the National Institute for Health and Care Research Doctoral Research Fellowship 302303: The views expressed are those of the author(s) and not necessarily those of the NIHR or the Department of Health and Social Care.

## Author contributions

Conceptualization, Formal analysis, Investigation, Methodology: 1^st^, 2^nd^, 4^th^

Project administration, Supervision, Writing-original draft: 1^st^, 2^nd^, 4th

Data curation, Writing-review & editing: all authors.

## Ethics declaration

The GenROC study received Research Ethics Committee (REC) approval on 15 December 2022 and Health Research Authority approval on 9 February 2023.

## Conflict of interest statement

None declared.

## Patient consent statement

All participants provided informed consent for interview as per GenROC study protocol

---

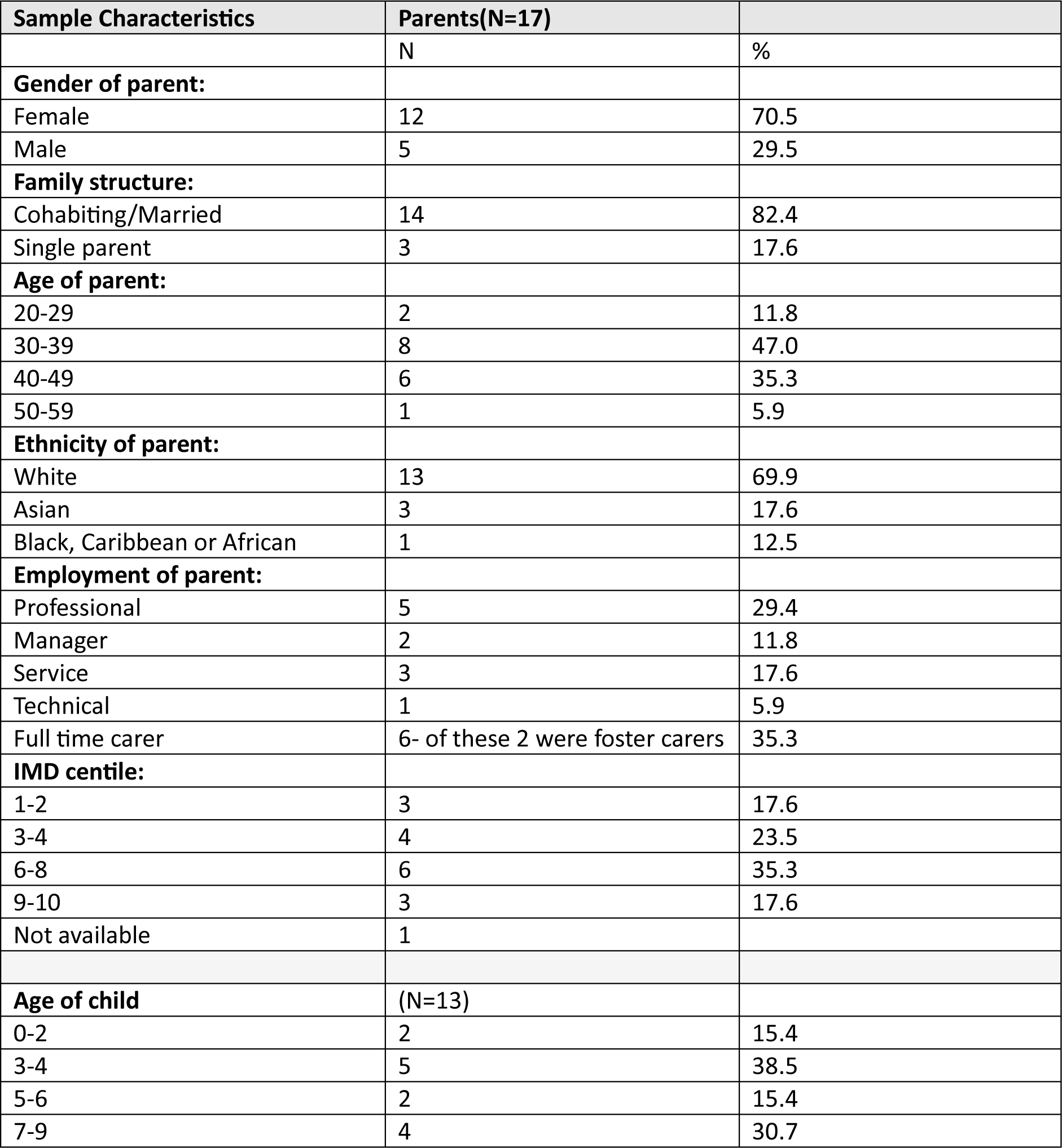

| Sample Characteristics | Parents(N=17) |  |
| --- | --- | --- |
|  | N | % |
| **Gender of parent:** |  |  |
| Female | 12 | 70.5 |
| Male | 5 | 29.5 |
| **Family structure:** |  |  |
| Cohabiting/Married | 14 | 82.4 |
| Single parent | 3 | 17.6 |
| **Age of parent:** |  |  |
| 20-29 | 2 | 11.8 |
| 30-39 | 8 | 47.0 |
| 40-49 | 6 | 35.3 |
| 50-59 | 1 | 5.9 |
| **Ethnicity of parent:** |  |  |
| White | 13 | 69.9 |
| Asian | 3 | 17.6 |
| Black, Caribbean or African | 1 | 12.5 |
| **Employment of parent:** |  |  |
| Professional | 5 | 29.4 |
| Manager | 2 | 11.8 |
| Service | 3 | 17.6 |
| Technical | 1 | 5.9 |
| Full time carer | 6- of these 2 were foster carers | 35.3 |
| **IMD centile:** |  |  |
| 1-2 | 3 | 17.6 |
| 3-4 | 4 | 23.5 |
| 6-8 | 6 | 35.3 |
| 9-10 | 3 | 17.6 |
| Not available | 1 |  |
| **Age of child** | (N=13) |  |
| 0-2 | 2 | 15.4 |
| 3-4 | 5 | 38.5 |
| 5-6 | 2 | 15.4 |
| 7-9 | 4 | 30.7 |

## Themes and subthemes

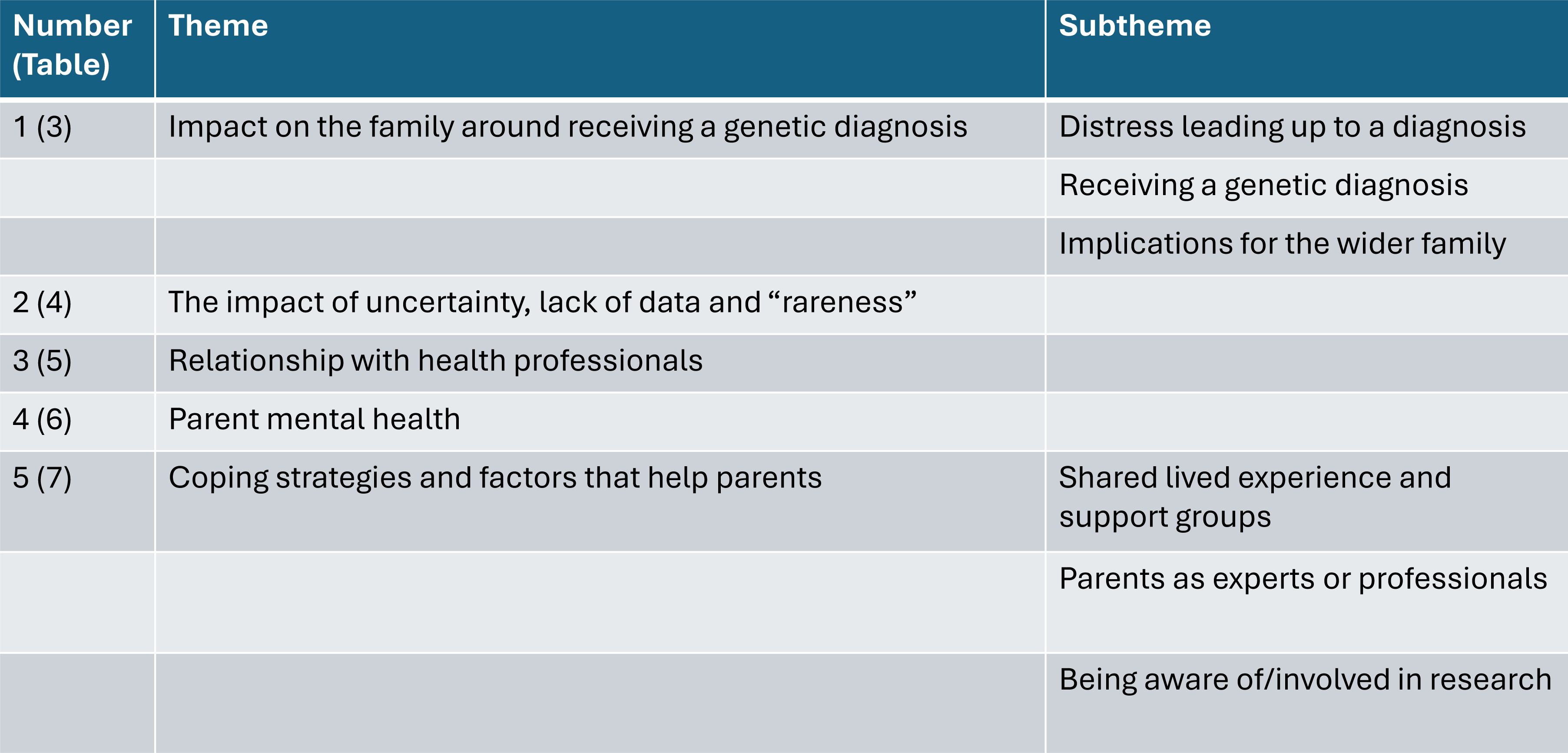

**Theme 1.** Impact on the family around receiving a genetic diagnosis.

| Quote number | Quote |
| --- | --- |
| 1 | "Then I went to the doctor with him, the paediatrician, and the first paediatrician was actually quite awful, don't think it was his fault, he was clearly having a bad day, and he was a locum, and he said, "I've never heard a mother complain to me that the baby doesn't cry before."(R10F) |
| 2 | I said, "I'm a bit worried he's having seizures." "No, no, babies do that it's fine, don't worry," and I am a paediatric nurse, I worked in a tertiary centre in a ward with lots of children with epilepsy, so it was like am I seeing what I think I'm seeing which are really subtle seizures, or am I being super paranoid because that's my work environment?"(R6F) |
| 3 | "first of all there was an appointment saying, "You need to come in and see us, we've found something, you need to come in and see us." So I don't like that, I prefer for them to say, "We have found this, and then come in and find out more information," rather than, "We found something," and it's like well what have you found? At least I can start talking to people about it and finding out bits."(R4F) |
| 4 | "Well we don't need that operation, but it's because her brain's not growing." As a parent getting the news that your child's brain isn't growing on the telephone from a junior doctor that was pretty devastating." (R13F) |
| 5 | "because in an appointment with [child], child is 100 miles an hour, 110 miles an hour, and it feels like when I come out of an appointment it feels like I've run a marathon. Because I'm trying to talk to the doctor and receive information, whilst stopping her cracking her head open." (R4F) |
| 6 | "Thanks for your letter, but it doesn't really tell me a huge amount, can I have the medical report? Can I have the genetics report?" (R4F) |
| 7 | "and we were like no, no, no it's fine, it's nothing, it's not... can't be that. Then she was doing it a bit more, and then we stayed in denial a little bit, because she wasn't doing anything else at that time, and then a friend said, "She does that weird thing that [son] used to do," and we were like, "No she doesn't, she doesn't." "(R6F) |

**Theme 2.** The impact of uncertainty, lack of data and “rareness”.

| Quote number | Quote |
| --- | --- |
| 8 | “ and then it’s like because he had a diagnosis, all these tests started stopping, it was like just part of his syndrome, and we were like but what if it’s not? And it just got more frustrating, even though we have the answers to what was wrong with him, it got more frustrating then because it were just like oh well he’s got a diagnosis now... they just blame it on his diagnosis all the time.”(R8F) |
| 9 | “ just going down a tunnel of I want to read up everything about this now, and just being up at early hours just reading through all these articles, and you should be sleeping, but you just want to do everything you can to help really. ... it’s difficult finding factual information that were credible sources.” (R5F) |
| 10 | "you want to try and know is what the future looks like a little bit, just so you can set your expectations and plan, because you don’t want to set yourself up for failure, but also you want some positivity in what you’re going to achieve.” (R5M) |
| 11 | “ they’re like, “Well what is it?” And I go, “It’s X,” and they go, “I’ve not heard of that,” and I’m like, “I know you haven’t, I told you it was one of the really rare conditions. ... whenever I explain what it is people tend to tell me, “Oh well I’ve got this person who’s got this, and it’s very similar,” and I’m like, “It’s not similar, it’s not the same.” (R7F) |
| 12 | “I think it’s a very challenging thing just the sheer universal luck of it all just it’s overwhelming to me, and makes me start thinking about very big topics that stress me out, and so I just don’t.” (R7F) |
| 13 | “”...just the finality of it. We’d got used to the idea that [daughter] was one and we were thinking we might get out of this... because some sorts of epilepsy they just grow out of it, and some sorts of epilepsy you can lead a normal life. But when we get that result it was... but it was just the seizures aren’t going to go away, and she’s going to have a severe learning disability. ” (R3M) |

**Theme 4:** Parent mental health.

| Quote number | Quote |
| --- | --- |
| 19 | I used to be a completely different person until I had [son] until... well it's not [son]'s fault, obviously it was all what happened, now I'm more one of them where I just sit alone don't I? I'd rather just go and sit upstairs, no telly on, and I just like to be on my own" (R8F) |
| 20 | " it's almost like they assume that you're being an over-anxious parent first, and then you have to prove them differently, it's not the other way round... and that can actually make you mad, it actually can. I know a lot of parents and carers that really have had very bad mental health from it." (R1F) |
| 21 | "Sometimes the behaviour can be so bad that it really is the biggest burden on family life... It means that we can't go out alone on the weekends with both of them, so you have to always have two adults. They're getting older, they're both mobile, they both are fully unaware of danger, they don't listen, they don't stop, they don't look back, they don't care if they've left you or not left you. They will run out in the road, they will walk off with strangers. They are not in tune with any of that, and if you've got one having a full on crazy meltdown that might last up to 40 minutes, and you've still got another one that you're trying to hold onto as well, you just can't do that on your own. So we can't do anything alone." (R6F) |
| 22 | "They're the moments, when you're at the beach and everyone's just sitting down and the kids are just playing nicely next to them, and the parents are all having a right old time, and they're the times where me and [partner] look at each other and think this is really... that's when we really feel it, because we can't just go and sit on the beach, the kids won't just sit next to us and play, they'll be gone. So it's... or [daughter] will be eating a handful of stones, or whatever. Yeah, so it's quite burdensome."(R6F) |

**Theme 5:** Coping strategies and factors that help parents.

| Quote number | Quote |
| --- | --- |
| 23 | “because they’re all so nice, and it’s like just one big family on there isn’t it? You don’t get judged for anything, and they are all... if you need any answers or anything you just put on there, and they’ll answer it for you. They are really... it is a really, really good group.” (R8F) |
| 24 | “then there’s times where you’re just you’re struggling with a problem, and you don’t think there’s an end in sight, and you’re thinking am I the only one in... is this... am I the only one with this problem? And then you reach out to people... has anyone had any success with coming... finding the source of the problem, or something that will alleviate it..” (R5F) |
| 25 | “because they’re having the exact same experience, and getting to understand beyond what the papers say about the spectrum of the disease, and what people go through, and what their children go through and how they present across the lifespan. There’s several adults in the group, so it gives us a little bit of a glimpse what the future might look like.” (R6F) |
| 26 | “They’re staring at you because it’s a window into their future, and because this is... people hate the unknown. So it’s almost like if they can see that you’re 16 and that you’re at school, and that you’ve passed your IT exams, that gives parents of younger kids hope.” (R10F) |
| 27 | so we went on to have another child, and it was a really big decision, and I don’t think we would have had the guts to do that if we hadn’t seen these other families on Facebook, and it’s like that showed us that it was possible... just seeing that there was all these other families on there where they’d gone on to have other children, and I think that influenced us quite a lot in terms of possibilities.” (R3M) |
| 28 | “But he wanted to know the co-ordinates so he could then go and research and... because he found it really useful to... was helpful for him so that he could then go and learn all that, and learn where the terminus was in the gene that meant that she didn’t make the protein, which meant that she then has these complications.”(R7F) |
| 29 | “you get the little code like P.3702 G to C or something, and they’re like what the hell does this mean? And so I’ll try and explain it, but apart from that everyone wants to know if it’s going to tell them how their kid’s going to be, and with X it won’t. So I tend to say that, and that happens quite a lot, because it’s really hard to explain.” (R3M) |
| 30 | “But I was floating around aimlessly for years, I didn’t know what to do because I had to give up my job, because I was a teacher, and then I was just caring for [daughter], and I’m not really very good at that kind of aspect. I would never have been a carer, it’s not something I would ever have chosen to do, so I wasn’t getting fulfilment out of just staying at home and caring for her... So I setup the website actually before I setup the charity.”(R13F) |
| 31 | “that’s also one of the reasons why I want to partake in research so that other people aren’t left in the dark , so we can keep others informed of it... So it’s just giving back in that sense I think.” (R5F) |

PPI Group reflections
| Quote number | Quote |
| --- | --- |
| 32 | "Thanks for sharing this. Wow, what a valuable piece of research! Quotes really resonate... Theme 4 is pretty difficult isn't it, although definitely rings true. As do the findings about coping strategies." |
| 33 | "Wow, I actually feel really emotional reading that, it's like we could be all those people and written those words ourselves! I think it will be really well received, and people will feel listened to. I guess it will have an impact on professionals too in terms on their delivery and care. I'm often trying to improve my delivery at work, when it's your day job it's easy to get so blase, but to the people you're dealing with it's their everything in that moment. " |
| 34 | "Honestly? I cried most of the way through reading this, hearing other parents struggles which have felt so familiar to our own. I know I shouldn't react so personally but I'm guessing that's what I'm here for!" |
| 35 | "The endless appointments you have with gp's/consultants/paediatricians etc who do not know anything about your child's condition - and that is fine, they can't be experts in everything - but have not even bothered to spend 5 minutes researching the basics of the condition before the appointment and proceed to 'google' it at the appointment, in front of you. It feels very disrespectful and really undermines parents' confidence in that professional directing your child's well-being, and it's clear you know so much more about your child's condition than they do. That leads on to the quote from parents saying that they have to fight to be heard - it often feels like you're carrying the weight of your child's wellbeing solely and you don't even have the support of health professionals to help you, it can feel quite overwhelming." |

## Notes

### Competing Interest Statement

The authors have declared no competing interest.

### Author Declarations

The GenROC study protocol was reviewed by the NHS Research Ethics Committee (Nottingham REC, United Kingdom). They gave ethical approval approval on 15 December 2022 and NHS Health Research Authority gave approval on 9 February 2023.

