## Supplementary material for "“They don’t know how to live with a child with these conditions, they can’t understand…”: The lived experiences of parenting a child with a genetic neurodevelopmental disorder": Topic guide

**The GENROC Study**: Improving the treatment of people with **GEN**etic **R**are disease: an **O**bservational **C**ohort study.

Working with families to increase understanding of rare genetic conditions in order to improve clinical care.

University of BRISTOL

**Topic guide for parent/carer participants – Focus groups**

GENROC

We will undertake in-depth discussions with parents/carers to understand their views and experiences of utilising the internet and social media to gain understanding of their child's genetic condition. Discussions will take place after the initial phase of online data collection. A sample of parents will be invited to take part. This topic guide details the areas that will be covered in the qualitative discussions. The interviewer may ask additional questions during the interview to clarify information. The questions may be minimally adapted throughout the process of interviewing as observations or alternate questions arise.

DATE: STUDY ID:

### 1. INTRODUCTIONS ☐

- **Introduce interviewer**
- **Introduce research**
    - Explain we are talking to parents/carers of children with genetic syndromes to understand how to use digital healthcare to improve our knowledge and the information available.
- **Thank participant** for taking part in the interview
- **Procedure**
    - We will be chatting to you with questions about your experience as a parent/carer in using online resources to help you understand your child's genetic condition
    - There are no right or wrong answers, I'm just interested in what you think
    - Options for breaks or stop at any time
    - Nothing they say will impact their child's treatment; will be using an audio-recorder
    - Explain I'll be taking notes for my benefit.
- **Confidentiality statement**
    - Everything we talk about will be confidential unless anything you say that worries me for your/your child's safety or for others
    - I'm **not** going to discuss any of our conversation with your child's clinician
- **Any questions?**

### 2. TURN ON DICTAPHONE & STATE IT'S ON ☐

### 3. STATE SIGNED CONSENT FORMS ☐

### 4. INTERVIEW TOPICS

### Setting the scene (10 mins)

1.**Can you tell me about your child's genetic diagnosis?**

PROMPTS: When was your child diagnosed? How was your child diagnosed? What prompted investigations?

2. **What information did you receive when your child was diagnosed?**

PROMPTS: Patient information leaflets? links to online resources? Links to social media groups? Patient support groups?

3. **Did you feel that you had enough information given to you?**

PROMPTS: How could this have been improved? Did you require follow up? What do you think should happen at follow up?

### Provision and acceptability of patient information & recruitment process (5 mins)

4.**What were your initial thoughts about the research study?**

Prompts: What did you think when you were told about it?

Feelings?

Worries?

Expectations?

**5.What did you think about the information you were given about the study?**

Prompts: What information did you get – oral and written (PIS)?

Did you read it?

Understand it?

Did it give you enough information/too much?

Was it appropriate for child's age?

Were there things you thought they had forgotten to include?

Was anything missing you wish you had known about the treatment or taking part?

**6.Did you discuss the study with anyone else e.g. your/their GP?**

Prompts: Clinical geneticist or paediatrician?

Are you getting any advice about your/their genetic condition from anyone else?

E.g. Online?

Other sources of information?

**7.What did you think about when deciding whether or not to take part in the study?**

Prompts: Feelings?

Expectations?

Worries?

Treatments offered?

What did you already know?

Did you think about the study in terms of helping other people or not?

### EXPERIENCE AND VIEWS OF DIGITAL HEALTHCARE/SOCIAL MEDIA (20 mins)

8.**Do you use the internet or social media?**

Prompts: give examples of social media such as facebook and whatsapp

9. **Do you use it to gain information about your child's genetic condition?**

Prompts: what would prompt you to go to the internet? what sort of information are you looking for? Can you given an example of when you have gone to the internet – good or bad? prompt a bit about both facts (e.g. about diagnosis/prognosis/treatments etc etc) and experiential information (other people's experiences, sharing their own etc)

When you go online do you consider reliability of information? Eg.is the information from a reputable scientific source; What would make you think a site is reliable? Unreliable? What do you consider to be reliable information?

10.**Are you a member of a social media group specific to your child's genetic condition?**

Prompts: How did you find out about the group? Is it a UK based group or international? Closed or open? If closed, what 'proof' is required? How many members? Is any sort of consent taken or implied? Are there any rules in the group about sharing information? Are researchers allowed in the group?

Are you a member of more than one group? (If so, Given opportunity to address above questions again) Why are you a member of more than one group? What are the differences? Are there conflicts between your experiences in the different groups?

11**. What is the main reason for your joining this group?**

Prompts: What prompted you to join? What do you see as the biggest benefit of being in the group? What is the downside (if any)? Are you seeking facts or the experience of support and not being alone.

11. **What are your views on the information that you have gained from these sites/groups**?

Prompts: Do you trust the information? Do you think it is reliable? Do you think it could be biased? Have you tried to use the information in a medical setting (expand) Do you think the data should be shared with doctors? If yes, how do you think this could be done? Do you think there are concerns around consent with regards to the data held by these groups? If yes, how could this be addressed in your opinion?

### PARENTAL VIEWS ON ONLINE PORTAL (15 mins)

First explain the idea of the online portal and what it would involve

13. **What do you think about the idea of a portal**?

Prompts: what do you think of this idea? what effects do you think this might have? 'how might people use it? Any possible benefits or harms? And how would this differ compared to social media? Would parents take the time to upload?

14. **Do you have any concerns about uploading data in this way**?

Prompts: Any concerns re identifiable data? Would you be happy to upload photos? What information would be important for parents to see first?

### OTHER ASPECTS OF GENROC (10 mins)

15. **Do you understand what data linkage means**?

Prompts: Did you read the information leaflet? Did you scan the QR code and read the further information/ watch the video? Did you understand what this meant? Do you understand that the linkage will continue in the future? What are your views on this? Should there have been different information? How so?

16. **We have included children in GENROC until they turn 16. What are your views on consent after 16?**

Prompts: What is your view on recontacting for consent at 16? Do you think this is important? Do you think families would find this a burden?

17.**What did you think of the questionnaires used in the study**?

Prompts: What were your thoughts on the questionnaire?

Did they ask the questions you wanted to answer?

Was there the right number of questionnaires?

What should a questionnaire ask?

18. **Do you have any other thoughts or anything else you would like to talk about?**

Prompts: About the study?

Taking part in research in general?

Are there any questions you would like to ask?

STOP DICTAPHONE
